# Social media use and exposure to pro-tobacco content: a comparison of sexual identity and gender in young adults in California

**DOI:** 10.1101/2024.09.19.24313969

**Authors:** Ollie Ganz, Nishi J. Gonsalves, Eugene M. Talbot, Scott I. Donaldson, Michelle Jeong, Jon-Patrick Allem

## Abstract

**Introduction:** Given the rapidly changing media landscape and tobacco marketplace, timely data on media consumption and exposure to pro-tobacco content across media channels among lesbian, gay, and bisexual (LGB) young adults is imperative for developing counter-messaging and public education campaigns for these individuals.

**Methods:** Using 2023 data from young adults in California, this study examined how social media use differed across media channels for heterosexual and LGB young adults, by gender identity. Exposure to pro-tobacco content across media channels between heterosexual and LGB young adults, by gender identity was also compared.

**Results:** Findings from a representative sample of young adults in California showed that more LGB young adults reported using Tumblr and fewer reported using Facebook and Snapchat, compared with heterosexual young adults, among both males and females. Additionally, social media use differed by gender identity. For example, use of Reddit was more common among LGB versus heterosexual females, but there were no differences by sexual identity among males. Exposure to tobacco marketing was more common among LGB females compared with heterosexual females, but this was not the case for males. Exposure to user-generated e-cigarette content, and self-reported visits to e-cigarette websites, were more common among LGB females compared to heterosexual females.

**Conclusions:** These findings highlight the importance of treating LGB individuals as a heterogeneous group. As such, anti-tobacco campaigns designed for LGB individuals that leverage social media will want to consider which social media platforms are most used among their target audience, ensuring maximum campaign reach.

## INTRODUCTION

Cigarette smoking prevalence among young adults in the United States (U.S.) has decreased in the past decade;^1^ however, cigarette and other tobacco use, including e-cigarettes, remains disproportionately high among lesbian, gay, and bisexual (LGB) young adults.^2,3^ While tobacco use disparities exist between heterosexual and LGB young adults, there is growing evidence of differences by sex and LGB subgroup,^2,4-6^ with LGB females reporting an especially high prevalence of cigarette smoking.^2,7^ Exposure to pro-tobacco content, which has been linked to subsequent tobacco use,^8,9^ may be driving elevated tobacco use among LGB young adults, as well as within-group disparities. Indeed, LGB young adults have reported greater recall of^10,11^ and receptivity^12,13^ to tobacco industry marketing, compared with their heterosexual counterparts. Mirroring patterns of tobacco use, young LGB females report disproportionately high recall of, and receptivity to, tobacco marketing.^11,12,14^

Studies suggest that tobacco marketing exposure via media channels differs between LGB and heterosexual individuals.^14,15^ For example, sexual minority males report greater odds of engagement with online tobacco marketing,^14^ and exposure to tobacco marketing via the internet, newspapers and magazines, and via TV and streaming services, compared with heterosexual males.^15^ Differing media use patterns may be partly responsible for these differences, with one 2013 study showing that LGB adults reported greater odds of having social media accounts, accessing Facebook daily, and frequent internet use, compared with heterosexual adults.^16^ These data, however, are over a decade old and the media landscape has changed dramatically, with the emergence of new social media platforms (e.g., TikTok).

Given the rapidly changing media landscape and tobacco marketplace, timely data on media consumption and exposure to pro-tobacco content across media channels, especially social media, among LGB young adults is imperative for developing public education campaigns for these individuals. Using 2023 data from young adults in California, this study examined how social media use differed across platforms for heterosexual and LGB young adults and compared exposure to pro-tobacco content across channels between these groups. This study stratified analyses by gender identity to understand within-group differences, which can be disguised when LGB individuals are treated as a monolith.^2^ Findings from this study can be used to inform anti-tobacco marketing education campaigns targeted at LGB young adults.

## METHODS

### Data Source

Young adults (18-24 years of age) living in California with access to an electronic device with the internet were recruited by YouGov, a research panel agency, to complete an online survey from 05/23/2023 to 06/27/2023. Participants (N=1500) were matched to a sampling frame based on age, gender, education, and race. The sampling frame was a politically representative modeled frame of U.S. adults based on the American Community Survey.^17^ The matched cases were weighted to the sampling frame using a propensity score matching procedure. Participants were sent a link directing them to an online survey. They provided informed consent, and completed survey items measuring their social media use, tobacco marketing exposures, tobacco use, and sociodemographic characteristics. This study was approved by the University of Southern California Institutional Review Board.

### Measures

#### Independent variables

Independent variables included gender (male or female) and sexual identity. Sexual identity was categorized through the question, “Which of the following best represents how you think of yourself?” Participants who selected “gay or lesbian,” “bisexual,” “something else,” or “I’m not sure yet,” were coded as LGB; if respondents selected “straight, that is, not gay or lesbian,” they were coded as heterosexual. Response options “don’t know what this question means” (n=1) and “prefer not to say” (n=36) were coded as missing. Additionally, respondents who selected “transgender female,” “transgender male,” “something else,” and “I’m not sure yet” to the gender identity question were excluded from analyses, due to the small sample size (n=54). As a result, the analytic sample was n=1404.

#### Dependent variables

Dependent variables included social media use and exposure to tobacco marketing. Social media use was captured by the question, “Which if any of the following social media sites do you use?” Respondents were presented with 10 social media sites: YouTube, Facebook, Reddit, Instagram, Twitter, TikTok, Pinterest, Snapchat, Tumblr, Twitch, and “none of the above.” For each site participants indicated using, they were asked how often they visited it. “Any use” was defined as using less often than once a month, monthly, weekly, once daily, or several times a day.

Offline tobacco advertising-industry sponsored exposure was defined as having seen an offline tobacco ad via billboards, posters, inside retail stores, or other in the past 30 days. Exposure to any user-generated e-cigarette content was defined as having seen content from: people they know in real life; from online friends they have not met in real life; random content pushed to their feed, but not from people they know; celebrities or social media influencers; or public health campaigns (e.g., Truth Initiative), in the past 30 days. To capture exposure to direct-to-consumer marketing, respondents reported if they signed up for any e-cigarette companies’ newsletters (e.g., email listserv) or received emails from any e-cigarette companies in the past 30 days. Respondents also reported visiting any e-cigarette company websites in their lifetime or in the past 30 days. To assess exposure to tobacco imagery in a film or series, respondents were first asked to select which films or series they had seen from a previously identified list containing e-cigarette imagery, based on ratings data from Nielsen Media Research.^18,19^ Those who selected yes to a given film or TV series were then asked, “did you see people using or talking about vaping e-cigarettes on [INSERT film/TV recall]?” Those who responded “yes” to this question for any of the films or series were categorized as having seen people using or talking about vaping e-cigarettes from select list of films and tv shows. Lastly, respondents who reported having ever seen an e-cigarette advertisement on social media were asked if they had seen one in the past 30 days. Those who answered “yes” were then asked to select which tobacco products they had seen advertised on social media in the past 30 days (i.e., cigarettes, older-generation e-cigarettes, disposable e-cigarettes, hookah, oral products).

### Analyses

Descriptive statistics for study sample characteristics, social media use, and tobacco marketing exposure were calculated and stratified by gender identity, comparing LGB to heterosexual individuals. Unweighted frequencies and weighted percentages were calculated for all tables. Significance was determined based on a p-value <0.05, corresponding to Rao-Scott adjusted chi-square test accounting for design effect. Analyses were weighted and conducted using SAS software V.9.4 (SAS Institute, Cary, North Carolina, USA) complex survey procedures.

## RESULTS

### Overall sample characteristics

A total of 22.6% of the sample was LGB. A majority of the sample was female (52.1%), Hispanic (49.2%) or non-Hispanic White (26.8%), age 21 or older (77.4%), and heterosexual (77.4%; **Supplemental Table 1**). A total of 13.92% of the sample reported past 30-day cigarette smoking and 19.1% reported past 30-day vaping. There were no significant differences in demographic characteristics or cigarette smoking and vaping between LGB and heterosexual/straight participants.

**Table 1.** Social Media Consumption by Sex and LGB status (N=1404)

|  | Males |  | p-value | Females |  | p-value |
| --- | --- | --- | --- | --- | --- | --- |
|  | Heterosexual<br>Unweighted n<br>(weighted %) | LGB<br>Unweighted n<br>(weighted %) |  | Heterosexual<br>Unweighted n<br>(weighted %) | LGB<br>Unweighted n<br>(weighted %) |  |
| YouTube |  |  |  |  |  |  |
| No use | 36 (7.13) | 9 (10.6) | 0.3022 | 57 (9.56) | 25 (8.63) | 0.6612 |
| Any use<sup>a</sup> | 423 (92.87) | 67 (89.4) |  | 535 (90.42) | 252 (91.37) |  |
| Facebook |  |  |  |  |  |  |
| No use | 255 (58.40) | 55 (73.5) | **0.0236** | 292 (48.59) | 166 (59.21) | **0.0063** |
| Any use<sup>a</sup> | 204 (41.60) | 21 (26.5) |  | 300 (51.41) | 111 (40.79) |  |
| Reddit |  |  |  |  |  |  |
| No use | 302 (64.99) | 45 (61.3) | 0.5643 | 492 (83.65) | 185 (68.69) | **<.0001** |
| Any use<sup>a</sup> | 157 (35.01) | 31 (38.7) |  | 100 (16.35) | 92 (31.31) |  |
| Instagram |  |  |  |  |  |  |
| No use | 95 (20.65) | 25 (32.8) | **0.0317** | 52 (8.89) | 31 (12.94) | 0.1068 |
| Any use<sup>a</sup> | 364 (79.35) | 51 (67.2) |  | 540 (91.11) | 246 (87.06) |  |
| Twitter |  |  |  |  |  |  |
| No use | 225 (50.46) | 38 (46.6) | 0.5698 | 348 (60.31) | 137 (50.09) | **0.0078** |
| Any use<sup>a</sup> | 234 (49.54) | 38 (53.4) |  | 244 (39.69) | 140 (49.91) |  |
| TikTok |  |  |  |  |  |  |
| No use | 216 (47.36) | 40 (51.1) | 0.5844 | 117 (19.42) | 52 (19.63) | 0.9473 |
| Any use<sup>a</sup> | 243 (52.64) | 36 (48.9) |  | 475 (80.58) | 225 (80.37) |  |
| Pinterest |  |  |  |  |  |  |
| No use | 413 (89.63) | 65 (86.5) | 0.4409 | 286 (49.45) | 117 (43.21) | 0.1092 |
| Any use<sup>a</sup> | 46 (10.37) | 11 (13.5) |  | 306 (50.55) | 160 (56.79) |  |
| Snapchat |  |  |  |  |  |  |
| No use | 253 (54.21) | 54 (70.7) | **0.0157** | 191 (33.14) | 129 (47.32) | **0.0002** |
| Any use<sup>a</sup> | 206 (45.79) | 22 (29.3) |  | 401 (66.86) | 148 (52.68) |  |
| Tumblr |  |  |  |  |  |  |
| No use | 446 (97.62) | 68 (86.9) | **<.0001** | 558 (94.83) | 224 (80.80) | **<.0001** |
| Any use<sup>a</sup> | 13 (2.38) | 8 (13.1) |  | 34 (5.17) | 53 (19.20) |  |
| Twitch |  |  |  |  |  |  |
| No use | 328 (72.04) | 58 (72.2) | 0.9837 | 529 (90.24) | 207 (74.54) | **<.0001** |
| Any use<sup>a</sup> | 131 (27.96) | 18 (27.8) |  | 63 (9.76) | 70 (25.46) |  |

NOTES: Unweighted frequencies and weighted percentages are reported; p-values correspond to Rao-Scott adjusted chi-square test (accounts for design effect); LGB=lesbian, gay, and bisexual
a: Values are collapsed (monthly, weekly, once daily, several times a day, and less often than once a month)

### Social media use

**Table 1** describes differences in social media consumption by sexual identity, stratified by gender identity. Findings show that a greater proportion of male participants identifying as heterosexual reported using Facebook (41.6% vs 26.5%, p=0.02), Instagram (79.3% vs 67.2%, p=0.03), and Snapchat (45.8% vs 29.3%, p=0.02) compared with LGB males. In contrast, a greater proportion of LGB males reported using Tumblr compared with heterosexual males (13.1% vs 2.4%, p<.0001). A greater proportion of heterosexual females reported using Facebook (51.4% vs 40.8%, p=0.006) and Snapchat (66.9% vs 52.7%, p<.001) compared with LGB females (Table 1). In contrast, LGB females reported disproportionate use of Reddit (31.3% vs 16.3%, p<.0001), Twitter (49.9% vs 39.7%, p=0.008), Tumblr (19.2% vs 5.2%, p<.0001), and Twitch (25.5% vs 9.8%, p<.0001) compared with heterosexual females.

### Exposure to tobacco marketing

**Table 2** describes differences in tobacco marketing exposure by sexual identity, stratified by gender identity. Exposure to offline tobacco marketing was more common among heterosexual versus LGB males (57.3% vs 43.6%, p=0.04). In contrast, exposure to offline tobacco marketing was more common among LGB versus heterosexual females (57.8 % vs 49.1%, p=0.0243).

**Table 2.**
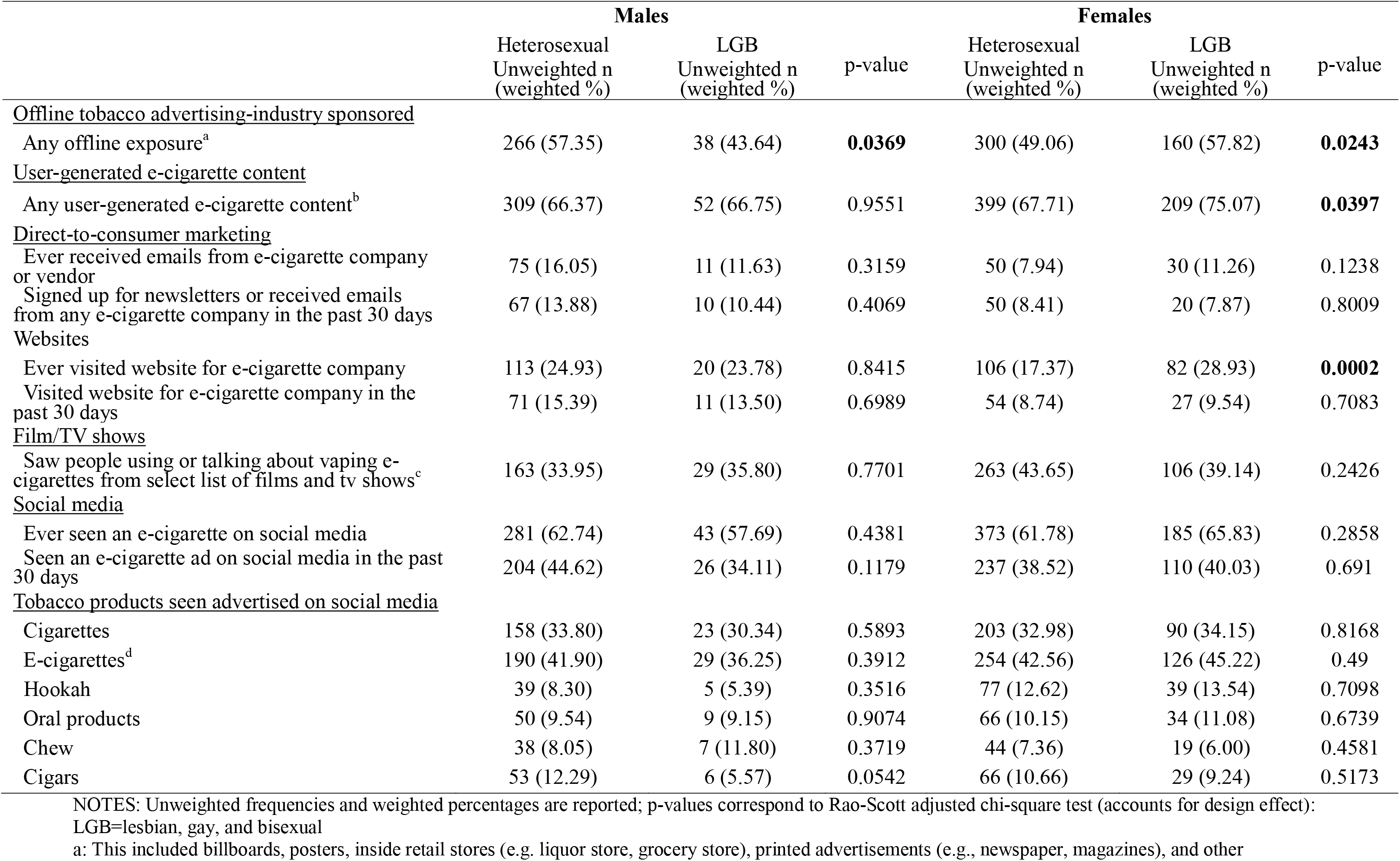
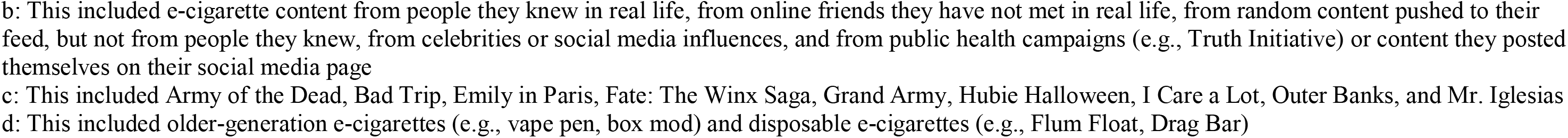
Exposure to Tobacco Marketing (N=1404)

|  | Males |  |  | Females |  |  |
| --- | --- | --- | --- | --- | --- | --- |
|  | Heterosexual<br>Unweighted n<br>(weighted %) | LGB<br>Unweighted n<br>(weighted %) | p-value | Heterosexual<br>Unweighted n<br>(weighted %) | LGB<br>Unweighted n<br>(weighted %) | p-value |
| <u>Offline tobacco advertising-industry sponsored</u> |  |  |  |  |  |  |
| Any offline exposure<sup>a</sup> | 266 (57.35) | 38 (43.64) | **0.0369** | 300 (49.06) | 160 (57.82) | **0.0243** |
| <u>User-generated e-cigarette content</u> |  |  |  |  |  |  |
| Any user-generated e-cigarette content<sup>b</sup> | 309 (66.37) | 52 (66.75) | 0.9551 | 399 (67.71) | 209 (75.07) | **0.0397** |
| <u>Direct-to-consumer marketing</u> |  |  |  |  |  |  |
| Ever received emails from e-cigarette company or vendor | 75 (16.05) | 11 (11.63) | 0.3159 | 50 (7.94) | 30 (11.26) | 0.1238 |
| Signed up for newsletters or received emails from any e-cigarette company in the past 30 days | 67 (13.88) | 10 (10.44) | 0.4069 | 50 (8.41) | 20 (7.87) | 0.8009 |
| <u>Websites</u> |  |  |  |  |  |  |
| Ever visited website for e-cigarette company | 113 (24.93) | 20 (23.78) | 0.8415 | 106 (17.37) | 82 (28.93) | **0.0002** |
| Visited website for e-cigarette company in the past 30 days | 71 (15.39) | 11 (13.50) | 0.6989 | 54 (8.74) | 27 (9.54) | 0.7083 |
| <u>Film/TV shows</u> |  |  |  |  |  |  |
| Saw people using or talking about vaping e-cigarettes from select list of films and tv shows<sup>c</sup> | 163 (33.95) | 29 (35.80) | 0.7701 | 263 (43.65) | 106 (39.14) | 0.2426 |
| <u>Social media</u> |  |  |  |  |  |  |
| Ever seen an e-cigarette on social media | 281 (62.74) | 43 (57.69) | 0.4381 | 373 (61.78) | 185 (65.83) | 0.2858 |
| Seen an e-cigarette ad on social media in the past 30 days | 204 (44.62) | 26 (34.11) | 0.1179 | 237 (38.52) | 110 (40.03) | 0.691 |
| <u>Tobacco products seen advertised on social media</u> |  |  |  |  |  |  |
| Cigarettes | 158 (33.80) | 23 (30.34) | 0.5893 | 203 (32.98) | 90 (34.15) | 0.8168 |
| E-cigarettes<sup>d</sup> | 190 (41.90) | 29 (36.25) | 0.3912 | 254 (42.56) | 126 (45.22) | 0.49 |
| Hookah | 39 (8.30) | 5 (5.39) | 0.3516 | 77 (12.62) | 39 (13.54) | 0.7098 |
| Oral products | 50 (9.54) | 9 (9.15) | 0.9074 | 66 (10.15) | 34 (11.08) | 0.6739 |
| Chew | 38 (8.05) | 7 (11.80) | 0.3719 | 44 (7.36) | 19 (6.00) | 0.4581 |
| Cigars | 53 (12.29) | 6 (5.57) | 0.0542 | 66 (10.66) | 29 (9.24) | 0.5173 |

NOTES: Unweighted frequencies and weighted percentages are reported; p-values correspond to Rao-Scott adjusted chi-square test (accounts for design effect):
LGB=lesbian, gay, and bisexual
a: This included billboards, posters, inside retail stores (e.g. liquor store, grocery store), printed advertisements (e.g., newspaper, magazines), and other
b: This included e-cigarette content from people they knew in real life, from online friends they have not met in real life, from random content pushed to their feed, but not from people they knew, from celebrities or social media influences, and from public health campaigns (e.g., Truth Initiative) or content they posted themselves on their social media page
c: This included Army of the Dead, Bad Trip, Emily in Paris, Fate: The Winx Saga, Grand Army, Hubie Halloween, I Care a Lot, Outer Banks, and Mr. Iglesias
d: This included older-generation e-cigarettes (e.g., vape pen, box mod) and disposable e-cigarettes (e.g., Flum Float, Drag Bar)

Exposure to user-generated e-cigarette content (75.1% vs 67.7%, p=0.04) and reporting having ever visited websites for e-cigarette companies (28.9% vs 17.4%, p<.001) were more common among LGB versus heterosexual females.

## DISCUSSION

Findings from a representative sample of young adults in California showed that social media use varied when LGB young adults were compared to heterosexual young adults. Social media use also differed by gender identity. Exposure to tobacco marketing also varied when LGB young adults were compared to heterosexual adults. Research has shown that pro-tobacco content was more common on certain social media platforms, including Instagram.^20^ Therefore, disproportionate use of these platforms by LGB individuals may increase disparities in exposure to pro-tobacco content, a well-established risk factor for tobacco use.^9^ Our findings show differences between LGB and heterosexual young adults in social media platform use, aligning with prior studies,^14,16^ but this study is the first to provide data on social media use patterns across a comprehensive platform list. We also found differences in tobacco marketing exposure. Most surprisingly, exposure to user-generated e-cigarette content and reporting having ever visited e-cigarette websites were more common among LGB females compared to heterosexual/straight females. Reasons for these observed differences among females remain unclear. While data show that adolescent females are more likely to follow e-cigarette influencers,^21^ and are also more impacted by social media influencers compared with males,^22^ this wouldn’t explain observed differences by sexual identity among females. However, given the findings of a recent meta-analysis showing an association between exposure to tobacco content on social media and tobacco use, including susceptibility to use tobacco among nontobacco users,^8^ it is troubling that the majority of LGB females were exposed to pro-tobacco user-generated content (e.g., posts on social media).

One prior study, using 2014-2015 Population Assessment of Tobacco and Health Study data, examined different ways in which LGB and heterosexual individuals were exposed to tobacco content (e.g., watching online videos about tobacco products).^14^ Findings showed that a greater proportion of sexual minority (i.e., LGB) females reported liking or following a tobacco brand on social media and sending a link about a tobacco brand on social media, compared with all other groups.^14^ However, prior research was limited to adolescents and only examined online tobacco marketing, disregarding offline channels (e.g., retail environment), or user-generated content (i.e., content not originating from a tobacco brand). The present study moves the literature forward by understanding within-group differences among LGB young adults, often hidden when LGB individuals are treated as a monolithic group in analyses.^2^

Several limitations should be considered in the current study. Data was collected from 2023 and may not generalize to other periods. LGB identities were measured as one category, precluding an examination of differences between these two groups, who have distinct tobacco use behaviors.^7^ Exposure to pro-tobacco content relied on self-report measures, which is subject to recall bias. Additionally, tobacco content presented to participants was not exhaustive. In other words, other pro-tobacco content will need to be considered in future research.

Despite these limitations, these findings have implications for public health education campaigns. Social media has been used previously to effectively reach the LGB community with anti-tobacco messaging.^23^ Indeed, the U.S. Food and Drug Administration’s “This Free Life” campaign, designed to reduce tobacco use among lesbian, gay, bisexual, transgender, and queer (LGBTQ+) young adults, used social media advertising and LGBTQ+ social media influencers to reach their target audience. Findings from the campaign’s evaluation demonstrated high reach among the target audience, but showed variation in campaign awareness by subgroup.^23^ For example, gay men had greater odds of ad awareness compared with lesbian/gay women, bisexual men, bisexual women and other sexual minority individuals.^23^ Findings from this study suggest that anti-tobacco social media campaigns designed for the LGB communities should leverage social media platforms most utilized by the specific subgroups that they intend to reach, or if there are notable disparities in campaign awareness by group, should adapt their social media strategy to focus on platforms used most frequently by those groups.

## Data Availability

Data and associated study materials will be made available upon request

## CONTRIBUTORS

OG: methodology, investigation, writing-original draft, writing-review & editing

NG: formal analysis; writing-original draft

ET: writing-original draft, writing-review & editing MJ: writing-original draft, writing-review & editing

SD: writing-original draft, writing-review & editing

JPA: conceptualization; funding acquisition; writing-original draft, writing-review & editing

## REFERENCES

1. Meza R, Cao P, Jeon J, Warner KE, Levy DT. Trends in US Adult Smoking Prevalence, 2011 to 2022. JAMA Health Forum. 2023;4(12):e234213–e234213.

2. Delahanty J, Ganz O, Hoffman L, Guillory J, Crankshaw E, Farrelly M. Tobacco use among lesbian, gay, bisexual and transgender young adults varies by sexual and gender identity. Drug and Alcohol Dependence. 2019;201:161–170.

3. Cornelius ME, Loretan CG, Jamal A, et al. Tobacco Product Use Among Adults - United States, 2021. MMWR Morbidity and Mortality Weekly Report. 2023;72(18):475–483. PMCID:PMC10168602.

4. Emory K, Kim Y, Buchting F, Vera L, Huang J, Emery SL. Intragroup Variance in Lesbian, Gay, and Bisexual Tobacco Use Behaviors: Evidence That Subgroups Matter, Notably Bisexual Women. Nicotine & Tobacco Research. 2016;18(6):1494–1501. PMCID:PMC5896797.

5. Romm KF, Huebner DM, Pratt-Chapman ML, et al. Disparities in traditional and alternative tobacco product use across sexual orientation groups of young adult men and women in the US. Substance Abuse. 2022;43(1):815–824. PMCID:PMC9372179.

6. Schuler MS, Collins RL. Sexual minority substance use disparities: Bisexual women at elevated risk relative to other sexual minority groups. Drug and Alcohol Dependence. 2020;206:107755. PMCID:PMC6980764.

7. Li J, Berg CJ, Weber AA, et al. Tobacco Use at the Intersection of Sex and Sexual Identity in the U.S., 2007-2020: A Meta-Analysis. American Journal of Preventive Medicine. 2021;60(3):415–424.

8. Donaldson SI, Dormanesh A, Perez C, Majmundar A, Allem JP. Association Between Exposure to Tobacco Content on Social Media and Tobacco Use: A Systematic Review and Meta-analysis. JAMA Pediatr. 2022;176(9):878–885. PMCID:PMC9274450.

9. U.S. Department of Health and Human Services. Preventing Tobacco Use Among Youth and Young Adults: A Report of the Surgeon General. Atlanta, GA: U.S. Department of Health and Human Services, Centers for Disease Control and Prevention, National Center for Chronic Disease Prevention and Health Promotion, Office on Smoking and Health. 2012.

10. Dilley JA, Spigner C, Boysun MJ, Dent CW, Pizacani BA. Does tobacco industry marketing excessively impact lesbian, gay and bisexual communities? Tobacco Control. 2008;17(6):385–390.

11. Tan ASL, Hanby EP, Sanders-Jackson A, Lee S, Viswanath K, Potter J. Inequities in tobacco advertising exposure among young adult sexual, racial and ethnic minorities: examining intersectionality of sexual orientation with race and ethnicity. Tobacco Control. 2021;30(1):84–93.

12. Ganz O, Krueger EA, Tan ASL, Talbot E, Delnevo CD, Cantrell J. Differences in Tobacco Advertising Receptivity Among Young Adults by Sexual Identity and Sex: Findings From the Population Assessment of Tobacco and Health Study. Annals of LGBTQ Public and Population Health. 2024;5(2):155–175.

13. Fallin A, Goodin A, Lee YO, Bennett K. Smoking characteristics among lesbian, gay, and bisexual adults. Preventive Medicine. 2015;74:123–130. PMCID:PMC4390536.

14. Soneji S, Knutzen KE, Tan ASL, et al. Online tobacco marketing among US adolescent sexual, gender, racial, and ethnic minorities. Addict Behav. 2019;95:189–196. PMCID:PMC6545129.

15. Azagba S, Shan L. Exposure to tobacco and e-cigarette advertisements by sexual identity status among high school students. Addict Behav. 2022;125:107165.

16. Seidenberg AB, Jo CL, Ribisl KM, et al. A National Study of Social Media, Television, Radio, and Internet Usage of Adults by Sexual Orientation and Smoking Status: Implications for Campaign Design. Int J Environ Res Public Health. 2017;14(4). PMCID:PMC5409650.

17. US Census Bureau. American Community Survey (ACS). https://www.census.gov/programs-surveys/acs. Published 2023. Accessed July 5, 2023.

18. Allem JP, Van Valkenburgh SP, Donaldson SI, Dormanesh A, Kelley TC, Rosenthal EL. E-cigarette imagery in Netflix scripted television and movies popular among young adults: A content analysis. Addict Behav Rep. 2022;16:100444. PMCID:PMC9253839.

19. Donaldson SI, La Capria K, Allem J-P. Recall of Netflix Scripted Content Known to Contain E-Cigarette-Related Imagery is Associated with Susceptibility to Use E-Cigarettes Among Young Adults. Subst Use Misuse. 1-5.

20. Vogel EA, Ranker LR, Harrell PT, et al. Characteristics of Adolescents’ and Young Adults’ Exposure to and Engagement with Nicotine and Tobacco Product Content on Social Media. Journal of Health Communication. 2024:1-11.

21. Lee J, Ouellette RR, Morean ME, Kong G. Adolescents and Young Adults Use of Social Media and Following of e-Cigarette Influencers. Subst Use Misuse. 2024:1-7.

22. Hudders L, De Jans S. Gender effects in influencer marketing: an experimental study on the efficacy of endorsements by same-vs. other-gender social media influencers on Instagram. International Journal of Advertising. 2022;41(1):128–149.

23. Guillory J, Crankshaw E, Farrelly MC, et al. LGBT young adults’ awareness of and receptivity to the This Free Life tobacco public education campaign. Tobacco Control. 2021;30(1):63–70.

