## Supplemental Table 1 for "Social media use and exposure to pro-tobacco content: a comparison of sexual identity and gender in young adults in California"

| Supplemental Table 1: Sample Characteristics by Sex and LGB status (N=1404) | | | | | | | |
| --- | --- | --- | --- | --- | --- | --- | --- |
|  | **Overall Sample**  Unweighted n (weighted %) | **Males** | |  | **Females** | |  |
|  |  | Heterosexual | LGB | p-value | Heterosexual | LGB | p-value |
|  |  | Unweighted n (weighted %) | Unweighted n (weighted %) |  | Unweighted n (weighted %) | Unweighted n (weighted %) |  |
| Sex |  |  |  |  |  |  |  |
| Male | 535 (52.11) | 459 (57.68) | 76 (33.07) |  |  |  |  |
| Female | 869 (47.89) |  |  |  | 592 (42.32) | 277 (66.93) |  |
| Race/Ethnicity |  |  |  |  |  |  |  |
| NH White^a^ | 421(26.78) | 157 (26.56) | 27 (29.84) | 0.1669 | 149 (23.93) | 88 (32.06) | 0.0954 |
| NH Black^a^ | 120 (6.87) | 45 (6.91) | 8 (7.23) |  | 42 (6.42) | 25 (7.52) |  |
| NH Asian^a^ | 174 (9.96) | 59 (10.18) | 13 (13.40) |  | 71 (9.42) | 31 (8.7) |  |
| NH Other^a^ | 125 (7.20) | 29 (5.74) | 10 (11.43) |  | 61 (8.22) | 25 (7.22) |  |
| Hispanic | 564 (49.20) | 169 (50.61) | 18 (38.10) |  | 269 (52.02) | 108 (44.43) |  |
| Age |  |  |  |  |  |  |  |
| Under 21 | 362 (22.64) | 98 (21.19) | 18 (25.82) | 0.4148 | 172 (23.73) | 74 (23.01) | 0.8226 |
| 21+ | 1042 (77.36) | 361 (78.81) | 58 (74.18) |  | 420 (76.23) | 203 (76.99) |  |
| Sexual identity |  |  |  |  |  |  |  |
| Heterosexual/Straight | 1051 (77.36) | 459 (100.00) |  |  | 592 (100.00) |  |  |
| LGB^b^ | 353 (22.64) |  | 76 (100.00) |  |  | 277 (100.00) |  |
| Tobacco use |  |  |  |  |  |  |  |
| Past 30-day cigarette smoking | 180 (13.92) | 88 (18.70) | 11 (12.66) | 0.2106 | 53 (9.16) | 28 (10.76) | 0.4803 |
| Past 30-day vaping | 263 (19.09) | 95 (20.43) | 13 (17.10) | 0.545 | 97 (16.59) | 58 (21.57) | 0.0945 |

NOTES: Unweighted frequencies and weighted percentages are reported; p-values correspond to Rao-Scott adjusted chi-square test (accounts for design effect) and indicate differences between heterosexual and LGB respondents; LGB=lesbian, gay, and bisexual

a NH: Non-Hispanic

b This included gay or lesbian, bisexual, something else, and I’m not sure yet
